# Biobank-scale exposome-wide risk factors in CAD and T2D: observational, predictive, and potentially causative evidence

**DOI:** 10.1101/2024.10.26.24316153

**Authors:** Sivateja Tangirala, Arjun K Manrai, John PA Ioannidis, Chirag J Patel

## Abstract

Cardiovascular disease and diabetes are intricately related and influenced by factors within the “exposome”. The exposome refers to a broad assessment of cumulative environmental exposures, including physical-chemical, biological, behavior, ecosystem, and social influences, experienced from conception throughout life, along with their dynamic interactions with the genome (Vermeulen et al. 2020). Distinguishing between correlational and potentially causative risk associations is challenging, especially at exposome scale. Here, we triangulate observational Exposome-Wide Association Study (*ExWAS*) evidence with “randomized” evidence for the exposome using mendelian randomization (MR) for almost 500 exposures. First, the *ExWAS* identified 144 significant factors for coronary artery disease (CAD) and 237 for type 2 diabetes (T2D), with 120 shared between both. These factors had modest predictive ability (variance explained) for both phenotypes. However, a genetic-based potentially causative relationship was deduced for only 14 factors in CAD and 16 in T2D, with seven implicated in both. Additionally, we found strong concordance of MR-validated findings between prevalent and incident disease associations (85.7% [12/14] for CAD and 87.5% [14/16] for T2D). Most correlational findings pertain to lifestyle factors (particularly diet), but social educational factors are more prominently highlighted among those with potentially causative support.

## Introduction

Type 2 diabetes (T2D) and coronary artery disease (CAD) are causatively related^1^ and carry a tremendous burden of disease worldwide. While genetics play a role, environmental factors – ranging from diet and pollution to social and behavioral factors – are thought to be key contributors to disease risk ^2^ , (Miller and Banbury Exposomic Consortium 2025)^3^.The totality of these external influences, collectively referred to as the exposome ^4^, encompasses diverse domains, including *social* (e.g., education and income), *behavior* (dietary and lifestyle), *physical-chemical* (e.g., nutrients and chemicals), *biological* (e.g., infection), and *ecosystem* (e.g. air pollution) variables. Understanding how these exposures contribute to disease remains a major scientific challenge.

Traditionally, epidemiological and toxicological studies have focused on a few pre-selected hypotheses, associating a limited number of exposures at a time. Hypothesis-driven studies are a way to test candidate relationships and, when conducted with methodological rigor and transparency, can yield robust and reproducible findings. However, such studies may be susceptible to certain limitations, including publication bias, confirmation bias, and a focus on pre-specified exposures, which can constrain the discovery of novel or unexpected associations—particularly in complex, multi-factorial environments such as the exposome where effect sizes may be moderate.

This narrow approach has led to inconsistent findings, potential false positives, and difficulty in replicating results across populations^5^. By contrast, genetic epidemiology has revolutionized risk factor discovery by systematically analyzing thousands of genetic variants in settings such as *genome-wide association studies (GWAS)*, improving reproducibility and replicability across a number of cohorts, and enhancing triangulation^6^. However, unlike genetic variants, exposure variables are time-varying, dynamic, and often highly correlated within themselves, making it difficult to disentangle their individual effects ^7^.

Importantly, considering only a handful of factors, leads to a fragmented and irreproducible literature of associations ^5,8^. Here, inspired by the genomics paradigm, we deploy an *exposome-wide association study* (ExWAS ^9,10^) framework, systematically screening a comprehensive array of exposures in an agnostic manner, while taking into account testing multiplicity ^8,11, 12^ . Second, by integrating ExWAS with Mendelian randomization (MR), we are among the first to bridge the gap between observational discovery and causal inference. This discovery-driven approach contrasts with existing MR studies that often focus on pre-specified exposures, enabling the identification of novel, unbiased relationships while maintaining the rigor of causal inference.

To “break” the correlation between exposures requires experimentation or “instrumental variables”, which serve as randomized proxies of exposure ^13^. However, the degree by which instrumental variables, or variables that can guide inference across the exposome is understudied^7^. Instrumental variables have been important in enhancing potentially causative determinations in other disciplines ^14^. With the advent of biobank data with both measured genotypes, exposures, and longitudinal phenotypic outcomes one can harness mendelian randomization (MR)^1, 15, 16^ as a way to scalably conduct “instrumental variable” analysis that hone in on identifying potentially independent causative relationships among exposures and phenotypic or disease outcomes.

Mendelian randomization (MR), which uses genetic variants as instrumental variables, offers an important strategy to reduce confounding and reverse causation in this setting. Nonetheless, MR can be challenged by violations of core assumptions, such as the exclusion restriction, particularly when applied to time-varying exposures ^17^ (PMID: 24681576).

We combine ExWAS and MR to identify the exposomic associations with T2D and CAD (Figure 1). First, we conducted an ExWAS systematically testing each of 495 individual factors of the exposome using a sample of 472,240 white European participants from the UK Biobank (UKB). and estimated its concordance with data from FinnGen, a biobank of 218,957 participants based in Finland. This approach allowed us to systematically evaluate which exposures had the strongest genetic-based evidence for potentially causative relationships and to distinguish between correlations and genuine risk factors.

**Figure 1.**
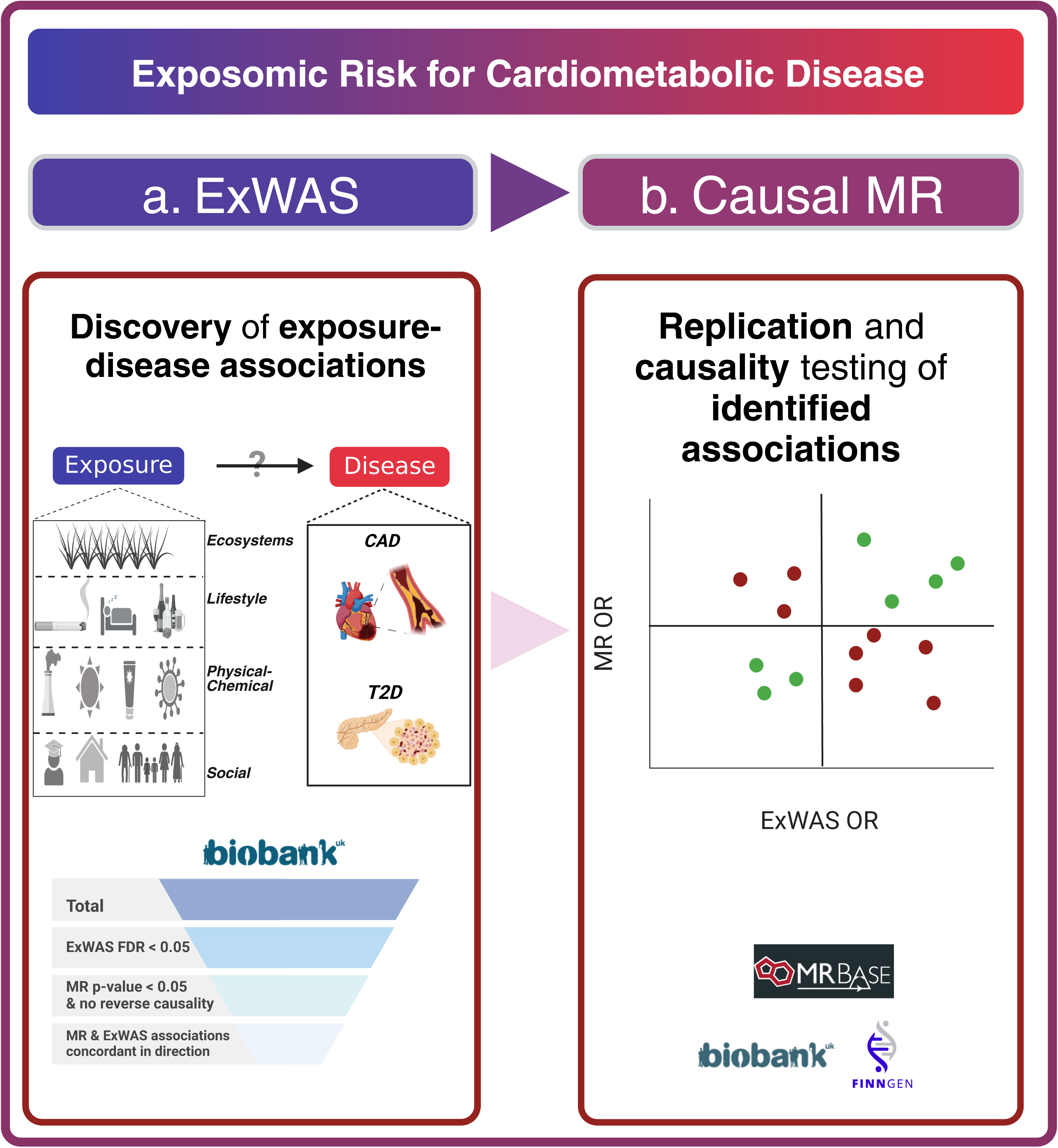
Schematic overview of analysis of exposome-wide risk architecture of T2D and CAD. This schematic diagram depicts our analytic workflow. (a) Exposure-wide association study (ExWAS) was performed to discover exposure-disease associations for T2D and CAD in the UK Biobank. (b) Two Sample bi-directional mendelian randomization (MR) was performed to replicate ExWAS-identified associations and test for potentially causative relationships. Briefly, genetic instruments for exposures were identified from the UK Biobank sample and tested on the FinnGen sample with respect to CAD and T2D

## Results

### Baseline characteristics

We report the baseline characteristics of clinical and demographic variables including age, sex, average household income, HbA1c, systolic blood pressure, diastolic blood pressure, BMI, LDL, HDL, triglycerides, total cholesterol, family history for CAD, family history for T2D, prevalent CAD, prevalent T2D, and smoking history of the UK Biobank (UKB) cohort participants in Table 1. Briefly, the average age of participants was 56.76, 54.48% were female participants, and ∼64% of participants had an average household income of less than 52,000 Euros. Out of the 472,240 European White individuals we utilize for our analysis, 7,426 have developed CAD and 12,050 have developed T2D (after baseline visit) while considering those who had already developed CAD (n=10,772) or T2D (n=17,303) at baseline separately. Additionally, out of the 218,957 participants in the FinnGen sample we utilize to evaluate our findings via two sample MR, 29,193 have developed T2D and 21,012 have developed CAD ^18^.

**Table 1.** Sample baseline characteristics. This table shows the mean (and standard deviation) or percentage of key clinical and demographic variables for the sample of White UKB participants that were primarily used for our analysis including age, sex, average household income, HbA1c, systolic blood pressure, diastolic blood pressure, BMI, LDL, HDL, triglycerides, total cholesterol, family history for CAD, family history for T2D, prevalent CAD, prevalent T2D, and smoking history.

| **Variable (units)** | **Label** | **Mean (SD) or Percentage** |
| --- | --- | --- |
| Age (years) | Age | 56.76 (8.028) |
| Sex (%) | Female | 54.48 |
| Sex (%) | Male | 45.52 |
| Avg. household income (%) | < 18,000 | 19.07 |
| Avg. household income (%) | 18,000 to 30,999 | 21.74 |
| Avg. household income (%) | 31,000 to 51,999 | 22.43 |
| Avg. household income (%) | 52,000 to 100,000 | 17.55 |
| Avg. household income (%) | > 100,000 | 4.65 |
| HbA1c (mmol/mol) | HbA1c | 35.96 (6.517) |
| Systolic pressure (mm Hg) | Systolic pressure | 139.9 (19.68) |
| Diastolic pressure (mm Hg) | Diastolic pressure | 82.16 (10.68) |
| BMI (kg/m2) | BMI | 27.41 (4.783) |
| LDL (mmol/L) | LDL | 3.57 (0.87) |
| HDL (mmol/L) | HDL | 1.45 (0.383) |
| Triglycerides (mmol/L) | Triglycerides | 1.75 (1.02) |
| Total cholesterol (mmol/L) | Total cholesterol | 5.71 (1.14) |
| Family history for CAD (%) | Family history for CAD | 39.8 |
| Family history for T2D (%) | Family history for T2D | 17.1 |
| Smoking (%) | Never | 53.94 |
| Smoking (%) | Previous | 35.55 |
| Smoking (%) | Current | 10.5 |
| Prevalent CAD (%) | Prevalent CAD | 2.28 |
| Prevalent T2D (%) | Prevalent T2D | 3.66 |

### Distribution of observational association sizes in incident CAD and T2D

We associated 495 exposures (Extended Data Table 1) with CAD and T2D. With FDR<0.05, we identified 144 significant exposures for CAD (Supplemental Table 1, Figure 2, Supplemental Results) and 237 significant exposures for T2D (Supplemental Table 2, Supplemental Figure 1, Supplemental Results). Association per exposomic category (as defined^4^) for each disease are in Extended Data Table 2. For CAD, exposomic category-wise representation among significant associations range from 2.33% (Ecosystems; exposomic factors pertaining to Ecosystems comprise 2.33% of significant associations) to 58.9% (Lifestyle). Similarly for T2D, category-wise representation among significant associations range from 2.59% (Ecosystems) to 52.8% (Lifestyle) .

**Figure 2.**
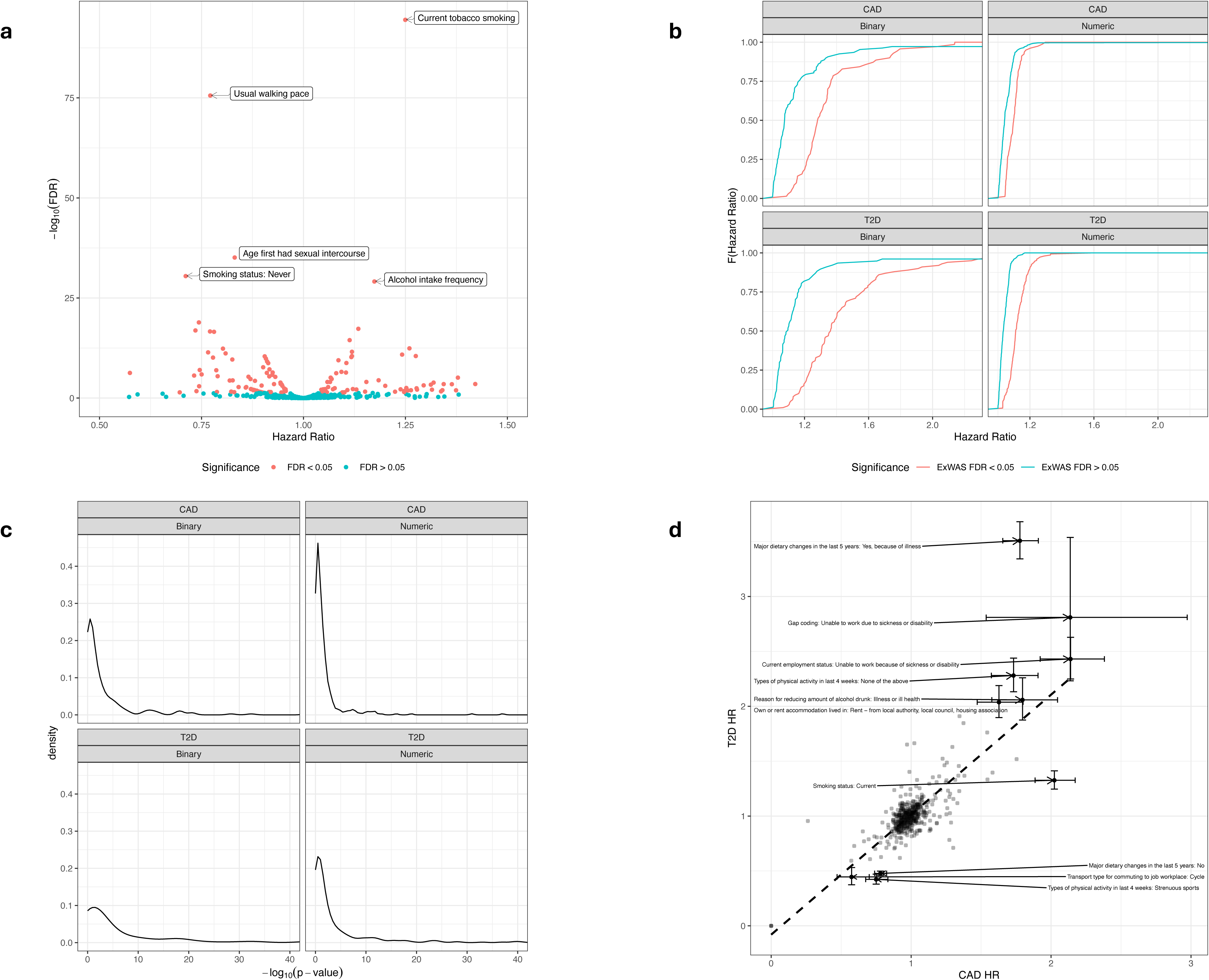
Exposomic architecture of CAD. This multipanel figure depicts exposome-wide findings for CAD. (a) **Volcano plot of ExWAS associations for CAD.** We visualize the FDR-corrected p-values on the negative log 10 scale versus hazard ratios of exposures. The red color indicates FDR < 0.05 significant associations and blue color indicates FDR > 0.05 associations. Top five associations are labeled. (b) **Distribution of hazard ratios of ExWAS associations for CAD and T2D stratified by significance.** The distribution of the hazard ratios (HR) computed from the absolute value of the regression beta coefficients for CAD and T2D are depicted with empirical cumulative distribution function plots and are colored by ExWAS significance. The distribution of HRs for exposures that are ExWAS FDR less than 0.05 significant are depicted in red. The distribution of HRs for exposures that are ExWAS FDR greater than 0.05 are depicted in blue. For the underlying dataset, n=472,240. (c) **Distribution of p-values of ExWAS and MR associations for CAD and T2D.** The distributions of all p-values derived from ExWAS for CAD and T2D are depicted with empirical cumulative distribution function plots. For the underlying dataset, n=472,240. (d) **Association size of CAD versus T2D for 495 exposures.** The hazard ratios (HR) of CAD versus T2D for 495 different exposures. Exposures that are FDR less than 5x10^-4^ significant for both CAD and T2D deviate and have HRs either less than 0.5 or greater than 2 for either disease are labeled and shown with error bars corresponding to the 95% confidence intervals for both CAD and T2D. The black dashed line on the scatterplot represents a linear regression line. For the underlying dataset, n=472,240.

**Table 2.**
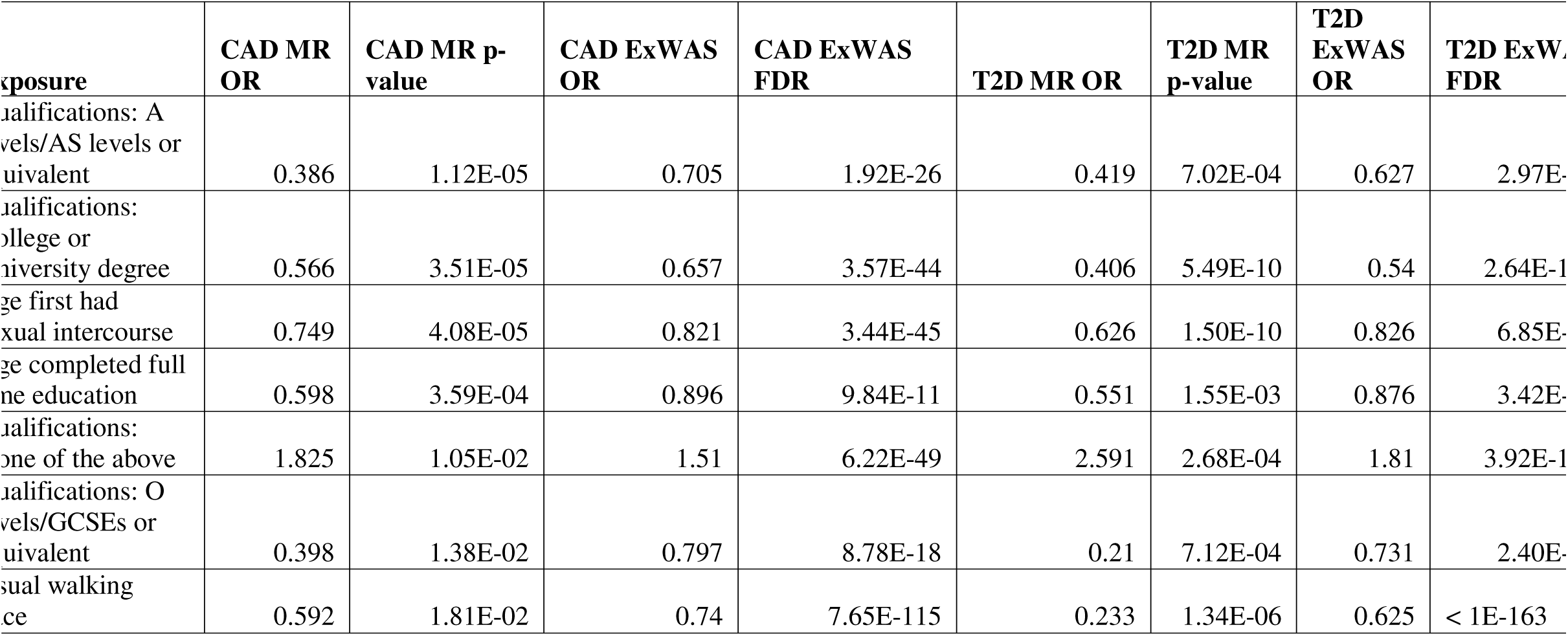
Exposomic variables significant in both MR and ExWAS and in both phenotypes. This table includes MR-derived odds ratios (ORs) and p-values and ExWAS-derived ORs and FDR-corrected p-values for the seven exposomic variables we found significant in both MR and ExWAS in both phenotypes.

For CAD, the IQR of HRs for significant continuous variables (for a 1 standard deviation [SD] change) that are protective factors is [0.89,0.94] and for risk factors it is [1.06, 1.11] while the IQR of HRs for significant binary variables that are protective factors is [0.75, 0.83] and for risk factors it is [1.25, 1.55]. For T2D, respectively, the IQR of HRs for significant continuous variables (for a 1 standard deviation [SD] change) are [0.86, 0.92] and [1.07, 1.13] and the IQR of HRs for significant binary variables are [0.64, 0.80] and [1.23, 1.53].

We found 120 significant exposures shared between T2D and CAD (119 of which are concordant and one discordant in direction of effect). Additionally, we found 117 exposures specific to T2D and 24 exposures specific to CAD. The Pearson correlation between the beta coefficients of CAD and T2D is 0.56 and 0.30 among continuous and binary exposure factors, respectively (Figure 2).

### Concordance of findings between prevalent and incident disease ExWAS

Additionally, we sought to assess the concordance of findings from ExWAS-identified associations for prevalent disease with those of incident disease. We associated 495 exposures with prevalent CAD and T2D. With FDR<0.05, we identified 232 significant exposures for prevalent CAD (Supplemental Table 3) and 309 significant exposures for prevalent T2D (Supplemental Table 4, Supplemental Results).

We found 137 significant exposures shared between prevalent and incident CAD (134 of which are concordant and 3 discordant in direction of effect). Furthermore, we found 62 exposures specific to prevalent CAD and 24 exposures specific to incident CAD. The Pearson correlation between the beta coefficients of prevalent and incident CAD is 0.53 and 0.38 among continuous and binary exposure factors, respectively (Supplemental Figure 2). Additionally, considering the prevalent disease ExWAS as our “diagnostic test”, we sought to compare the concordance of the prevalent CAD ExWAS with incident CAD ExWAS by computing the sensitivity and specificity of the prevalent CAD ExWAS test. We estimate the sensitivity to be 84.8% and specificity to be 65.2% (Supplemental Table 5).

Similarly, we compare the concordance between exposure associations for prevalent and incident T2D. We found 189 significant exposures shared between prevalent and incident T2D (184 of which are concordant and 5 discordant in direction of effect). Furthermore, we found 48 exposures specific to prevalent T2D and 24 exposures specific to incident T2D. The Pearson correlation between the beta coefficients of prevalent and incident T2D is 0.79 and 0.75 among continuous and binary exposure factors, respectively. We estimate the sensitivity and specificity of the prevalent T2D ExWAS test relative to incident T2D ExWAS to be 88.5% and 61.9%, respectively (Supplemental Table 6).

### MR-based assessment of observational ExWAS associations

We sought genetic-based evidence for potentially causative relationships and performed bi-directional MR between each exposure-outcome pair for which Genome-wide Association Study [GWAS] summary statistics were available (for 123/144 (85.4%) of CAD FDR significant associations and 182/237 (76.8%) of T2D FDR significant associations) to test whether the observational associations from the ExWAS are potentially causative. To enhance comparability, we use odds ratios (ORs) of exposures computed from logistic regressions after assessing the concordance in estimates between hazard ratios computed from Cox proportional hazard models and odds ratios computed from logistic regressions (Supplemental Figures 3 and 4). We visualize the concordance (and/or discordance) between ExWAS and MR ORs among these exposure-disease pairs in Figure 3 for CAD (and Supplemental Figure 5 for T2D).

**Figure 3.**
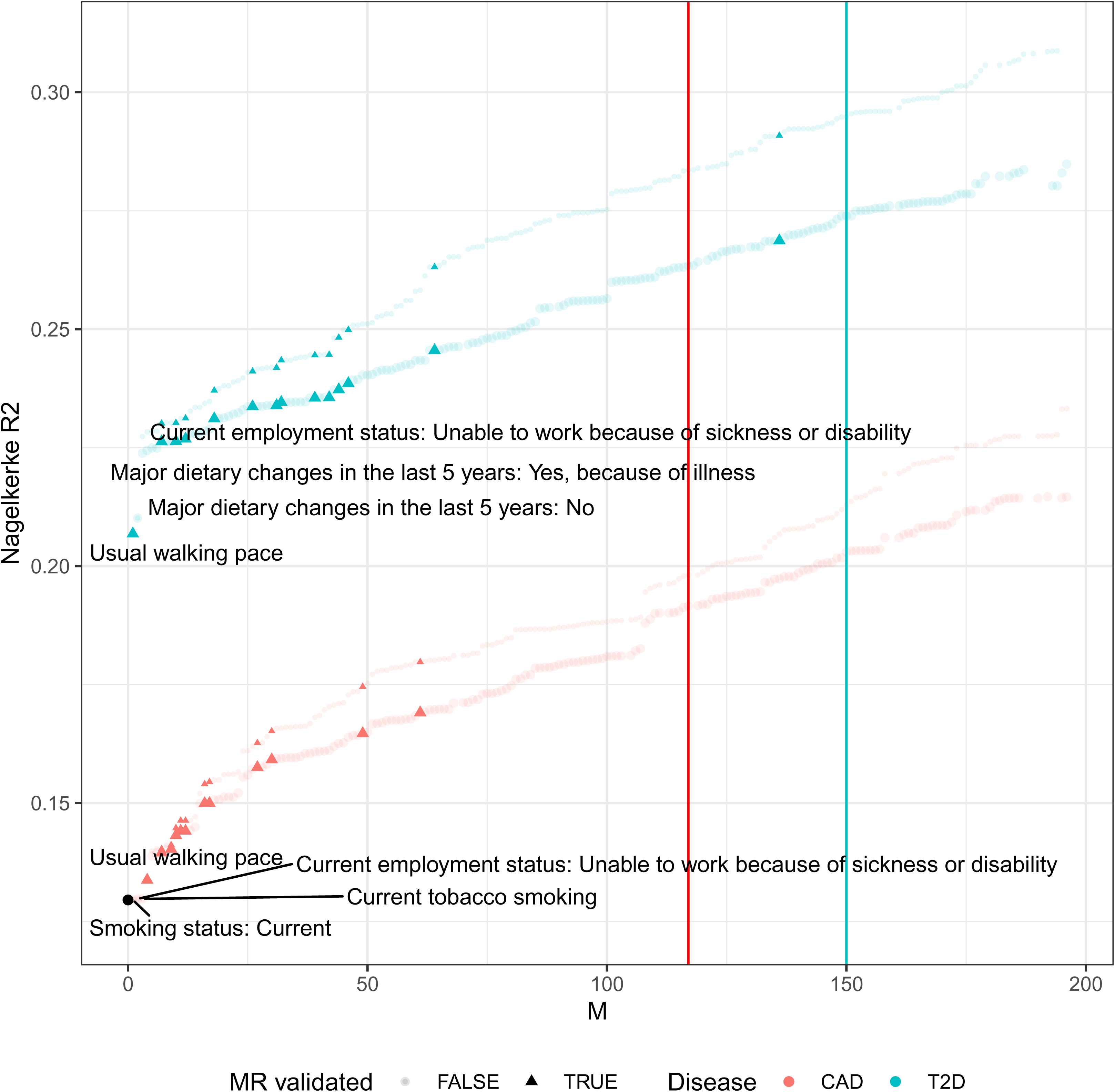
ExWAS association size versus MR causative estimate for CAD. The odds ratios (OR) computed from ExWAS versus the OR computed from MR for 260 different exposures in association with CAD. The blue color indicates evidence for potential reverse causative relationship and the red color indicates no evidence for potential reverse causative relationships. Triangles indicate associations with significant MR p-values. Horizontal and vertical black lines demarcate OR of 1 (null association). For the underlying datasets, n=472,240 for the UK Biobank sample and n = 218,957 for the FinnGen sample.

**Figure 4.**
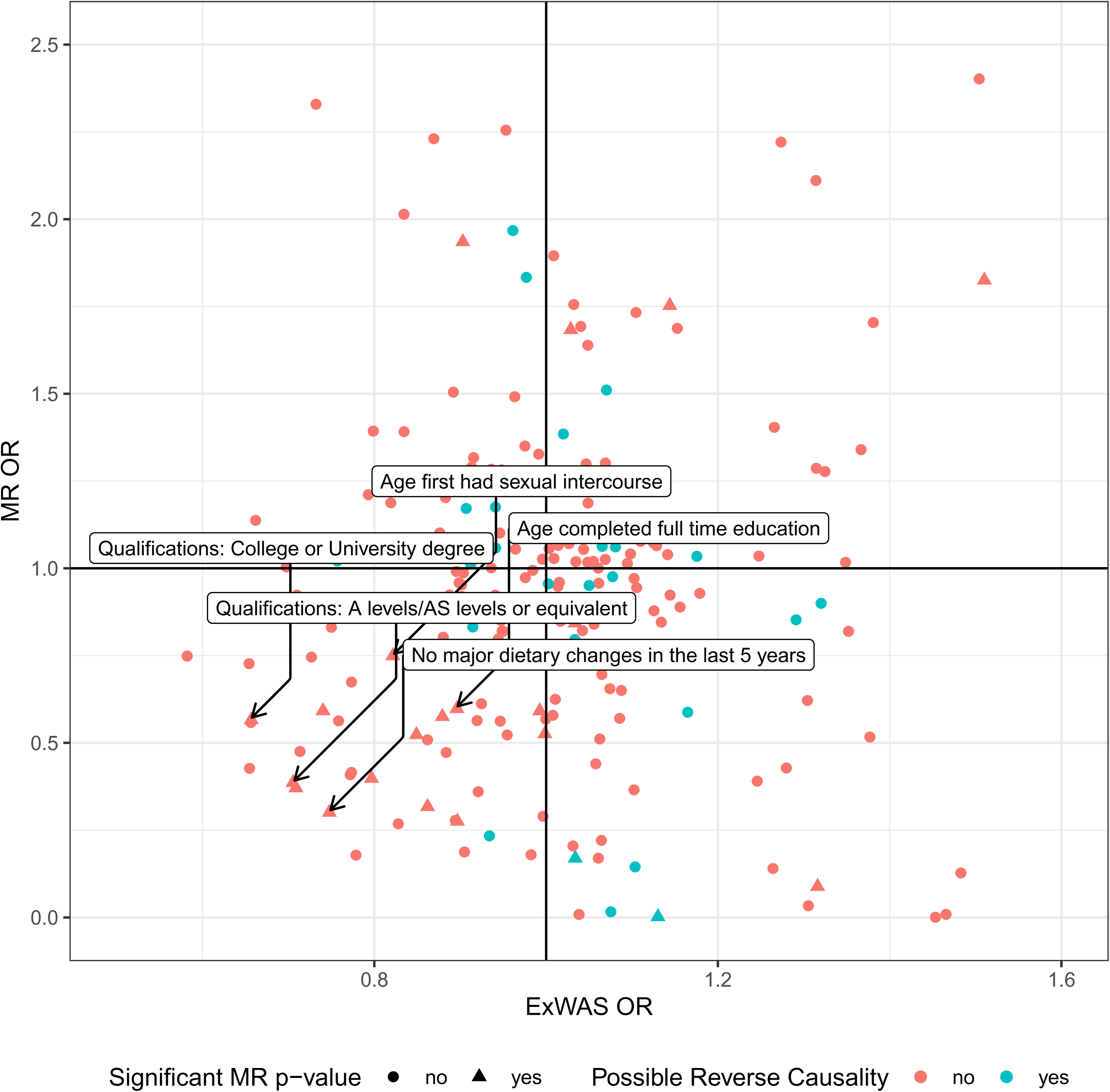
Visualizing the change in variance explained by step-wise addition of exposomic variables to baseline model of demographic and clinical risk factors compared to exposure-specific univariate models. We visualize the Nagelkerke R^2^ of each model constructed by step-wise addition of exposomic factors in descending order of FDR significance to the original baseline model comprised of demographic, genetic risk score (for CAD and T2D) and clinical (BMI for T2D) factors. Black dot indicates variance explained by the baseline model. Top exposures (by FDR) for the multivariable analysis are labeled. We also overlay the univariate (exposure) model results. We visualize the cumulative exposure-specific Nagelkerke R^2^ as we iterate through each univariate (exposure) model in descending order of FDR significance with smaller point size for both CAD and T2D, separately.

We identified 14 associations as being nominally significant (p-value less than 0.05) in forward MR and *not* p-value significant in the reverse direction in CAD while also being found to be concordant in direction to the corresponding ExWAS associations. The IQR of the ORs for CAD among MR-validated associations was [0.38, 0.58] and [1.79, 6.15] for protective and risk factors, respectively. The IQR of ORs for T2D among MR-validated associations was [0.25, 0.63] and [1.86, 2.38] for protective and risk factors, respectively. We also formally tested the difference in effect sizes (absolute value of beta estimates) of MR-validated factors and non-MR-validated factors with the Mann-Whitney test. MR-validated factors had stronger effects than non-MR-validated ones for CAD (Mann-Whitney statistic (W) = 571, p-value = 0.0044) and possibly also for T2D (W= 935, p-value = 0.021).

The MR results for CAD and T2D are made available in Supplemental Tables 7 and 8, respectively. Additionally, we found 12 out of the 14 total (85.7%) prevalent ExWAS-identified associations validated by MR to be the same as the MR-validated ones for incident ExWAS for CAD. Similarly, we found 14 out of the 16 (87.5%) total prevalent ExWAS-identified associations validated by MR to be the same as the MR-validated ones for incident ExWAS for T2D.

We describe the associations that were not only identified as significant from the ExWAS but also found to be significant (p-value less than 0.05) from the MR analysis for CAD (while also concordant in direction). The top 3 MR-validated protective factor associations (in order of increasing MR p-value) were having the educational qualification of A levels/AS levels or equivalent (MR OR: 0.39, MR p-value: 1.12x10^-5^, ExWAS OR: 0.71, ExWAS FDR: 1.92x10^-26^), having the educational qualification of college or university degree (MR OR: 0.57, MR p-value: 3.51 x10^-5^, ExWAS OR: 0.66, ExWAS FDR: 3.57x10^-44^), and the age participant first had sexual intercourse (MR OR: 0.75, MR p-value: 4.08x10^-5^, ExWAS OR: 0.82, ExWAS FDR: 3.44x10^-45^). The top 3 MR-validated risk factors were Townsend deprivation index at recruitment (MR OR: 1.75, MR p-value: 5.86x10^-3^, ExWAS OR: 1.14, ExWAS FDR: 9.23x10^-20^), having no educational or other professional qualifications (e.g. nursing, teaching) (MR OR: 1.83, MR p-value: 0.0105, ExWAS OR: 1.51, ExWAS FDR: 6.22x10^-49^), and no type of physical activity in the last four weeks (MR OR: 10.5, MR p-value: 0.0313, ExWAS OR: 2.07, ExWAS FDR: 2.37x10^-73^).

Additionally, MR results across the full suite of MR methods tested for are made available in Supplemental Tables 9 and 10. Among the top ExWAS-determined hits with MR-confirmed associations for CAD, we observed consistent effect directionality across all MR methods for most (9/14) exposures. Furthermore, among these 9 exposures with consistently concordant direction, statistical significance at p < 0.069 was seen in at least 3 of the 5 MR models for 2 of them, but none of the 9 had formally statistically significant results in all 5 models. For example, “Usual walking pace” showed directionally concordant inverse associations with CAD risk across methods (ranging from -1.408 [simple mode] to -0.525 [IVW]) and “Age first had sexual intercourse” showed (estimates ranging from -0.521 [simple mode] to -0.157 [MR Egger]). In contrast, for a subset of 5 exposures, such as “Age completed full-time education” and “Qualifications: A levels/AS levels or equivalent”, and “Qualifications: A levels/AS levels or equivalent,” the majority of MR methods yielded consistent directionality, but MR-Egger results showed discordance. These findings highlight the robustness of our primary findings while emphasizing the need for cautious interpretation in cases where sensitivity analyses indicate potential pleiotropy.

We also report the proportion of MR-validated exposures by exposomic category for CAD. Among social factors, 7.87% (7/89) were MR-validated and among lifestyle factors 5.3% (7/132) were MR-validated. More specifically, among education factors, 62.5% (5/8) were MR-validated and among dietary factors only 4.1% (3/73) were MR-validated. None were MR-validated among ecosystems and physical-chemical factors. Similarly, we report the proportion of MR-validated exposures by exposomic category for T2D. Among social factors, 6.9% (6/87) were MR-validated and among lifestyle factors 6.8% (9/132) were MR-validated. More specifically, among education factors, 62.5% (5/8) were MR-validated and among dietary factors only 4.1% (3/73) were MR-validated. 4.8% (1/21) were MR-validated among physical-chemical factors and none were MR-validated among ecosystems factors.

Next, we sought to better understand the concordance of our ExWAS test with respect to MR by computing the sensitivity and specificity of ExWAS (considering ExWAS as our “diagnostic test”). We extended our MR analysis to compute associations for all ExWAS tested disease-exposure pairs for which GWAS summary statistics were available [518 (52.3% of 990 total ExWAS-tested disease-exposure pairs)]. More specifically, we estimated the degree to which findings were both ExWAS significant and MR significant, or sensitivity (77.8% for CAD and 94.1% for T2D). Additionally, we estimated the degree to which ExWAS non-significant associations were also found to be MR non-significant, or specificity (37.3 % for CAD and 26.3% for T2D) (Extended Data Tables 3 and 4).

We observed a difference in the sensitivity and specificity of ExWAS between the lifestyle and social exposomic variable categories. For lifestyle factors, we estimate 70% sensitivity for CAD and 75% for T2D and 31.3% specificity for CAD and 21.4% for T2D [Supplemental Tables 11 and 12]. For social factors, the sensitivity was 87.5% for CAD and 85.7% for T2D and specificity was 45% for CAD and 34.6% for T2D [Supplemental Tables 13 and 14]. The correlation between ExWAS and MR beta estimates were 0.146 and -0.0271 among binary and continuous variables, respectively.

We identified 7 associations (5 education-related variables and 2 lifestyle factors [age first had sexual intercourse and usual walking pace]) in both T2D and CAD (Table 2) with MR. Here, we report two types of OR, both those obtained through ExWAS (a conventional OR) and those from the MR. These variables included age first had sexual intercourse, educational qualification of college or university degree, usual walking pace, having no educational or other professional qualifications (e.g. nursing, teaching), educational qualification of A levels /AS levels or equivalent, educational qualification of O levels/GCSEs or equivalent, age completed full time education. For example, we identified the educational qualification of college or university degree to be among the top associations for both T2D (MR OR: 0.407, MR p-value: 5.49x10^-10^, ExWAS OR: 0.54, ExWAS FDR: 2.64x10^-137^) and CAD (MR OR: 0.566, MR p-value: 3.51 x10^-5^, ExWAS OR: 0.657, ExWAS FDR: 3.57x10^-44^). Furthermore, we found all of the factors to be protective (OR less than 1) “other professional qualifications (e.g. nursing, teaching)” which we found to be a risk conferring factor for both CAD (MR OR: 1.83, MR p-value: 0.0105, ExWAS OR: 1.51, ExWAS FDR: 6.22x10^-49^) and T2D (MR OR: 2.59, MR p-value: 2.68x10^-4^, ExWAS OR: 1.81, ExWAS FDR: 3.92x10^-163^).

### Assessing the variance of CAD and T2Ds explained by exposomic, demographic, genetic and clinical risk factors

For incident CAD, we find that 42.3% of the variance explained by the exposome and demographics (Extended Data Table 5) can be explained by exposomics alone (e.g., for T2D, exposures alone explained 8% of the variance, while exposures plus demographics explained 18.8% of the variance). Additionally, 15.8% of the variance explained by the model including MR validated exposures and demographics is explained by MR-validated exposures alone.

Moreover, we observed how the variance explained by the models change after inclusion of T2D PRS and BMI. Exposomic variables alone account for 60.5% of the variance explained by the full model. Similarly, MR validated exposures alone account for 18.9% of the variance explained by the full model. We describe the T2D results in the Supplemental Results section.

Additionally, we computed the Nagelkerke R^2^ from logistic regression models that were run by serially adding exposomic factors to the previous model in order of increasing FDR-corrected p-values in addition to the baseline demographic and clinical risk factors (Figure 4). The IQR of the absolute value of pairwise correlations of the 196 exposures was [0.01, 0.08] (Supplemental Figure 6). For CAD and T2D, the Nagelkerke R^2^ for models ceased to increase when incorporating beyond the top 175 exposures by FDR-corrected p-values.

### Contextualization of MR-validated exposures in risk for CAD and T2D with established clinical risk factors

Finally, we run clinical risk factor models for CAD and T2D consisting of age, sex, family history, BMI, systolic blood pressure, diastolic blood pressure, HbA1c, LDL, HDL, triglycerides, total cholesterol, and smoking history (inspired by prior literature ^19, 20^ ). We compare the effects of these factors with those of seven MR-validated exposures commonly implicated in CAD and T2D (Supplemental Table 15). The magnitude of the effects of the MR-validated exposures matches or sometimes even exceeds the magnitude of the effects of established clinical risk factors.

## Discussion

Our analysis integrates ExWAS and MR methods to characterize the exposomic architecture of factors associated with CAD and T2D, made possible by use of biobank data, such as UK Biobank and FinnGen. Using ExWAS, we identified 144 (out of 495 [29.1%]) and 237 factors (out of 495 [47.9%]) for CAD and T2D, respectively. The larger number of factors identified for T2D may reflect, at least in part, the larger power available for T2D which was more common than CAD in the biobank sample. Overall, statistically significant ExWAS associations for these major outcomes seem to be too numerous. This may also explain the plethora of significant associations published in the epidemiological literature, where exposures are usually tested one at a time or a few at a time^21^.

For both CAD and T2D the magnitude of the effect sizes of ExWAS-identified factors was modest. Furthermore, the vast majority of them showed no genetic-based evidence for a potentially causative relationship on MR assessment. MR is an approach that uses genetic variants as “instruments” to test for a potentially causative association between exposures and disease ^15^. From our MR analysis, the validated factors tended to have somewhat larger effect sizes than those seen with observational correlation analyses in ExWAS. Only 14 (9.72%) and 16 (6.75%) of ExWAS findings were concordant with MR findings for the two phenotypes, CAD and T2D, respectively. We observed higher (genetic-based) potentially causative relationship rates among social and more specifically, education-related factors than for lifestyle factors, even though the latter accounted for the vast majority of ExWAS associations. While ExWAS may be sensitive when a large database like the UK Biobank is analyzed, specificity is quite low for T2D and CAD relative to MR, if MR is seen as the standard.

The strong presence of education-related variables among the apparently potentially causative ones is in line with other studies, including MR-based ones. For example, Tillmann et al. found that educational attainment is potentially causative and protective (OR: 0.67 for a 1 SD increase in higher education roughly corresponding to 3.6 years of additional schooling^22^) risk factor for coronary artery disease in the Coronary Artery Disease Genetics Consortium (CARDIoGRAMplusC4D), also suggesting increased levels of education have a protective effect ^22^. Additionally, we identified some factors suggestive of behavior including the age first had sexual intercourse in association with CAD and T2D risk and this is also in accordance with prior evidence^23^. Focused and intense “lifestyle” interventions that target weight loss for individuals with elevated glucose have been developed that have causatively led to decreases in T2D incidence^24^. Interventions may need to be tailored and tested targeting social and, in particular, educational factors. Such interventions would require a broader community outlook. Life course studies that examine education early in life may also be needed.

Our MR analyses incorporated a comprehensive suite of sensitivity methods beyond IVW, enabling us to assess the robustness of our findings and evaluate the potential for horizontal pleiotropy, which is a major threat to MR studies. Across the top exposures identified for CAD, we observed directionally concordant estimates across multiple MR approaches, including MR-Egger, weighted median, and mode-based methods. Although p-values were often attenuated in these sensitivity approaches—likely reflecting their lower statistical power relative to IVW—the consistent effect direction reinforces the plausibility of potentially causal relationships.

MR-Egger estimates for some exposures showed directional discordance relative to other methods. This highlights the potential for pleiotropy, underscoring the importance of cautious interpretation and the need for future studies with larger sample sizes and stronger instruments to confirm these findings, and potential multi-variable approaches. Current mega multi-phenotype studies have shown a vast potential for pleiotropy (Levin et al. 2025^25^), adding to the threat.

We also explicitly assessed potential sample overlap between the exposure and outcome GWAS used in our analyses. To the best of our knowledge, there is no overlap between the cohorts generating exposure GWAS summary statistics (primarily from the Neale Lab^26^ and MRC IEU OpenGWAS^27^ ) and the FinnGen cohort used for outcome data, minimizing the risk of inflated type I error.

Additionally, reverse MR analyses were conducted as a sensitivity test to assess potential reverse causative relationships, rather than to infer causative effects from disease to exposures. While such analyses can provide insight into the directionality of associations, their interpretation is limited by potential issues such as instrument strength and biological plausibility. Our reverse MR findings were used to support the primary forward MR results by evaluating the likelihood of reverse causative relationship.

Prior work have investigated potentially causative relationships for CAD and T2D through MR analyses. For example, the study by Georgiou et al. ^28^ focuses on specific lifestyle factors, such as alcohol consumption and physical activity, in relation to CAD, while the systematic review by Yuan et al.^29^ synthesizes MR findings for a range of pre-identified exposures relevant to T2D.

These studies provide valuable insights into targeted hypotheses but differ fundamentally from our discovery-driven, agnostic approach. Specifically, prerequisites for these studies include documented hypothesis between an exposure and an outcome. Such targeting of a specific hypothesis may require consideration of other types of evidence, but it may succumb to confirmation and other cognitive biases that perpetuate preferred narratives.

In contrast, our study employs an exposome-wide association study (ExWAS), which systematically identifying potential exposure-outcome relationships across a broad range of domains before validating these relationships with MR. The comprehensive, data-driven methodology is analogous to GWAS in its agnostic screening of a large number of exposures, extending the utility of MR beyond its traditional use in hypothesis-driven studies. By integrating ExWAS with MR, our approach enables the identification of both well-characterized and novel exposures that may have been overlooked in pre-specified analyses. For example, while some of the MR-validated associations we identified align with previously known findings, such as the protective role of educational attainment for CAD, our ExWAS uncovered associations with behavioral traits, such as age at first sexual intercourse, which were not previously highlighted in systematic MR investigations of CAD and T2D.

Our study systematically quantifies the low concordance between ExWAS findings and MR- validated associations for CAD and T2D. This underscores and estimates the extent of false positives in observational analyses, particularly for lifestyle and dietary factors, where we observed minimal potentially genetic causative evidence. Notably, our results demonstrate disproportionately high rates among education-related factors relative to other domains, a finding consistent with prior MR studies but newly quantified across a broad exposomic landscape. This systematic perspective reveals a disjoint between correlational and potentially causative structures for many lifestyle factors, while providing updated evidence for social and education-related exposures.

However, while our study highlights the potential of agnostic discovery-driven methods, it also underscores the challenges in establishing potentially causative evidence for many exposures, particularly lifestyle and dietary factors when using genetics as the instrument.

Among many lifestyle dietary factors, none survived MR scrutiny for potentially causative relationships in our data. While some factors may not have been detected in MR analyses due to low power, the overall pattern suggests a disjoint between correlational and potentially causative structures of different types of factors associated with these two major disease phenotypes. With rare exceptions, detected associations with lifestyle factors (most prominent among them being dietary factors) may reflect plain correlations rather than potentially causative relationships.

This is congruent with the evidence that while millions of published articles present or discuss observational associations with specific dietary and other related lifestyle factors, the far smaller corpus of interventional randomized trials relevant to direct effect of these exposures yields mostly null results^30, 31^. Moreover, other investigators using MR methods in large genetic consortia for CAD, T2D and ischemic stroke found no potentially causative evidence for any of the dietary factors that they tested and found only one potentially causative association (with fat) for heart failure^32^. One alternative model to consider includes the relationships between the wide array of exposures interrogated. For example, one hypothesis is that dietary and other lifestyle exposures are shaped by education-related factors, but do not by themselves cause cardiometabolic disease. Other hypotheses may be built around the correlation of lifestyle factors with weight which is a major causative risk factor for T2D and CAD (e.g., obesity may coexist with specific lifestyle patterns or even shape lifestyle features, but without many of these lifestyle features causing T2D and CAD by themselves). Future studies may test more complex potentially causative patterns from diverse directed acyclic graphs and mediation patterns among these multiple exposures.

Overall, we provide an overview of the currently measured exposomic risk architecture shared between T2D and CAD. Specifically, we can explain up to ∼8% and ∼12% in CAD and T2D outcomes respectively with current biobank scale measurements leaving much to be ascribed to newer measurements and instruments^4^. However, we find low concordance between MR and ExWAS for both CAD and T2D. Restricting the variables to just MR-implicated findings, we find that these variables explain 2 and 3% of variation, in CAD and T2D respectively. Although a crude comparison, heritability ranges have been reported from 30-50%, leaving much to be explained with the exposures considered in this study. The main reason for lack of concordance includes confounding with ExWAS findings reflecting false positive associations. Additionally, we suggest potential classes of exposures to assay for a larger number of participants and with greater resolution in the future. For example, the physical-chemical exposome (e.g., chemical pollutants, etc.) needs to be more comprehensively assayed at biobank scale^33^.

We also note limitations of our study. First, given the small sample size of individuals of non-White ethnic groups, we expect the results to have high uncertainty in MR approaches; therefore, we have excluded other ethnic groups from our analysis. A systematic review of MR studies for CAD suggests that direction of effects of modifiable risk factors tends to be similar in different ethnic groups, but differences in magnitude of effects may not be uncommon^34^. Second, MR methods may also have bias and flaws, and they are not a perfect gold standard by any means.

MR relies on three core assumptions: that the genetic instruments are strongly associated with the exposure (relevance), are independent of confounders of the exposure-outcome relationship (independence), and influence the outcome exclusively through the exposure (exclusion restriction). Violations of these assumptions can introduce bias and invalidate causal inferences. Furthermore, MR strictly identifies associations that are consistent with lifelong genetically influenced variations in exposure, and thus is particularly stringent in isolating causally robust pathways. Its assumptions—including genetic variants having a stable, lifelong impact on exposure, no pleiotropy, and no reverse causation—make MR an excellent choice primarily for robust causal signals with large effect sizes. However, exposures of relevance may have small or modest effects and may act only in specific periods of the lifespan.

In this study, many of the exposures analyzed are complex, distal variables within the exposome, rather than molecular traits that may be more proximal to genetic variation. As such, the genetic instruments used may be more susceptible to horizontal pleiotropy, where variants influence the outcome through alternative pathways. While we employed sensitivity analyses—including MR-Egger regression and the weighted median estimator—to assess the robustness of our findings and detect potential pleiotropy, these methods have limited power, particularly in settings with weak instruments or complex traits. Moreover, limited power may lead to false-negative potentially causative^35^ findings, but some biases may actually also create larger potentially causative effects^36^. Third, it is unclear if we have addressed confounding for all domains of exposures as there is no consensus on which adjustment factors to use for the different domains; future work could look at optimizing the difference between observational and MR associations as a function of covariate selection. Results may vary depending on what analytical choices are made^37^. Another important limitation of this study is potential under-ascertainment of T2D, as primary care data, medication use, and laboratory measures were not incorporated. These sources present challenges for time-to-event modeling, including unclear diagnosis timing, reverse causative relationships, and limited data availability. To maintain consistency and ensure temporal clarity, both prevalent and incident T2D analyses were based solely on hospital records and self-reported diagnoses. Future studies integrating comprehensive primary care data may improve case detection and further validate these findings. Additionally, while we adjusted for a key socioeconomic variable, socioeconomic status is multifaceted, and residual confounding may remain.

Finally, prevalent disease outcome data coupled with genetic variant data as instrumental variables for MR may suffice to identify exposures, especially when working with cohorts without sufficient longitudinal outcome follow-up data. This is evidenced by the high degree of commonality of MR-validated exposures between prevalent and incident disease ExWAS- identified associations (85.7% [12 out of 14 total] for CAD and 87.5% [14 out 16 total] for T2D). However, prevalent and incident outcome data may each have its sets of distinct biases.

In conclusion, MR-validated factors for CAD and T2D are far fewer than the rich range of factors identified by ExWAS. Potentially causative relationships also seem more commonly documented for educational factors, while lifestyle factors have many ExWAS signals, but these rarely have potentially causative MR support. Additional similar analyses in more cohorts with relevant information would be useful to expand evidence on the robustness of these findings.

## Online Methods

### Study population

The UK Biobank cohort is a prospective cohort including over 500,000 participants of ages 40-69 during recruitment from 2006-2010^38^. Differences between the UK Biobank cohort individuals and the general UK population were studied by Fry et al. in order to better understand sampling uncertainty^39^. Their study suggested that nonparticipants are more likely to be male, younger, and live in more socioeconomically deprived areas than UK Biobank participants ^39^. Information regarding how the UK Biobank data is maintained and validated can be found at https://biobank.ndph.ox.ac.uk/∼bbdatan/Data_cleaning_overall_doc_showcase_v1.pdf.

The National Research Ethics Service Committee North West Multi-Centre Haydock has approved the UKB cohort research and written informed consent to participate in the study was provided by all participants^40^. We analyzed *n* = 472,240 European White individuals (out of total *N* = 502,628 participants in the cohort). The other top three ethnicities represented (by sample size) in the UKB cohort (Indian, Caribbean, and African) were low in sample size (Indian n = 5,951, Caribbean n = 4,517, African n = 3,394) when integrating exposure data and therefore we may not have adequate power for detection of associations. Given the small sample size of individuals of non-white ethnic groups, we expect the results to have high uncertainty and thus the correlations would be weaker; therefore, we have excluded other ethnic groups from our analysis. Approval for the use of this data was approved by the UK Biobank (project ID: 22881). The Harvard internal review board (IRB) deemed the research as non-human subjects research (IRB: IRB16-2145). Formal consent was obtained by the UK Biobank (https:// biobank.ctsu.ox.ac.uk/ukb/ukb/docs/Consent.pdf).

### Disease outcomes

In our study, we investigate two major cardiometabolic incident outcomes in the UKB, including coronary artery disease (CAD) and type 2 diabetes as ascertained by using ICD-10 and self-reported disease status information (Supplemental Table 16). We identify cases of CAD and T2D as ones that occur after the baseline visit (incident cases) while considering individuals who have the disease at baseline (prevalent cases) separately. Primary care data, prescription records, and laboratory measures (e.g., HbA1c) were not included due to limitations in event timing, risk of reverse causative relationships, and incomplete data coverage.

### Categorizing ecosystems, lifestyle, social, and physical-chemical exposures

We considered 538 total variables of which we had an adequate number of complete cases to investigate 495 “exposures” that can be categorized (as per Vermeulen et al.^4^) as ecosystems, lifestyle, social, and physical-chemical factors throughout the paper. We utilize the data for these exposures collected during the participants’ baseline visits (2006-2010). These 495 exposures spanned 17 UK Biobank-defined categories (e.g., education, smoking, greenspace and coastal proximity, sun exposure, estimated nutrients yesterday) (Extended Data Table 1).

We averaged quantitative factors (e.g., infectious antigens [25 exposures]) across measurements from multiple visits. For exposures that did not have many observations in subsequent instances, we used only the data from the baseline visit (first instance of measurement collected during 2006–2010) (e.g., environmental factors from the estimated nutrients yesterday category). We also performed rank-based inverse normal transformation (INT) of these factors, as was suggested by Millard et al.^41^, to address non-normality and improve the comparability of effect estimates across exposures with differing distributions. This transformation ensures that all continuous exposure variables are on the same scale, reduces the influence of outliers and allows for more robust association testing in data-driven epidemiological analyses such as ExWAS . For categorical variables (which were also collected from multiple visits of a participant to the assessment center), we used data from the baseline visit (first instance of measurement collected during 2006–2010) as this contained the highest number of observations. Additionally, categorical variables with multiple levels were converted to sets of binary variables where each such variable indicates whether a participant has a given value of this variable (as was suggested by Millard et al.^41^). Ordinal categorical environmental factor variables were analyzed by treating such variables as continuous variables and real-valued quantitative environmental factor variables were scaled.

### Data-driven identification of exposure-disease associations

We used Cox proportional hazard models to associate each of the 495 factors and CAD (coronary artery disease), T2D (type 2 diabetes) [individually], while adjusting for sex, age, assessment center, ethnicity, average total household income after tax, and 40 genetic principal components (PCs) (computed and provided by the UK Biobank). PCs were included to account for subtle population structure that could correlate with non-genetic exposures, minimizing potential confounding due to population stratification.We adjust resulting p-values of associations for multiple comparisons using the false discovery rate (FDR) approach^42^. We report hazard ratios and FDR-adjusted *p*-values for the associations. Additionally, we run logistic regression to enhance comparison for downstream analyses (i.e. MR).

### Concordance between observational and MR-based exposure-disease associations

For each exposure-outcome pair, we conducted two-sample Mendelian randomization (MR) analyses using the full suite of methods available in the TwoSampleMR R package ^27^. The primary method used was inverse variance weighted (IVW) MR, which assumes that all genetic instruments are valid and provides the most precise estimates when this assumption holds. To assess the robustness of our findings and evaluate the potential for horizontal pleiotropy, we additionally applied complementary sensitivity analyses including MR-Egger regression, weighted median estimator, simple mode and weighted mode estimators, and the Wald ratio methods. These analyses enable us to assess consistency of effect direction and identify potential violations of MR assumptions, particularly related to horizontal pleiotropy.

Furthermore, we performed MR for all associations for which there were available GWAS summary statistics for the corresponding exposures (n = 292 exposures). We recognize that some exposures, including air pollution, are measured on the geospatial level, and it does not make sense to execute a MR on these^43^. We exclude such exposures and related findings from our MR analysis. Additionally, it is important to note that educational attainment was analyzed using different categories to align ExWAS exposures with GWAS summary data available for the UK Biobank and public resources (e.g., Neale lab^26^). We derived these measures from the PHESANT pipeline^41^, ensuring consistency across observational and MR analyses. Moreover, we used GWAS summary statistics for identifying instruments for each exposure from GWAS summary statistics generated by the Neale Lab^26^ and MRC IEU OpenGWAS ^27^ and made freely accessible on the MR-Base platform ^27^. We use the TwoSampleMR R package ^27^ to test the identified instruments for each exposure for each of the two outcomes using summary statistics of GWAS from the FinnGEN cohort (https://www.finngen.fi/en). To the best of our knowledge, there is no overlap between cohort utilized by the Neale Lab^26^ and MRC IEU OpenGWAS ^27^ to derive GWAS summary statistics for a given exposure and the FinnGen cohort^18^. Summary statistics from FinnGen were adjusted by sex, age, genotyping batch, and ten principal components. We utilize the FinnGen sample of 218,957 participants to validate our findings via two sample MR. Among these participants, 29,193 have developed T2D and 21,012 have developed CAD ^18^. Additionally, we used the following thresholds to ascertain genetic instruments including p-value < 5x10^-8^, linkage disequilibrium (LD) R^2^: 0.001, and clumping distance of 10000 kb. We report potentially causative genetic-based estimates computed using the inverse-variance weighting (IVW) method between each exposure-disease pair.

As a sensitivity analysis to assess potential reverse causative relationship, we performed reverse Mendelian randomization (MR), using genetic instruments for type 2 diabetes (T2D) and coronary artery disease (CAD) as exposures and the previously analyzed exposures as outcomes. Reverse MR was conducted using two-sample inverse variance weighted (IVW) MR as the primary approach. These analyses were not interpreted as evidence of causative effects but served to assess the possibility that genetic liability for disease influenced the exposures.

### Assessing the variance of CAD and T2D explained by exposomic and demographic risk factors

We contextualize the variance (computed as Nagelkerke R^2^) explained by validated exposomic factors with demographic risk factors (Extended Data Table 5). Briefly, for each incident disease we run two major sets of models: a) regressing incident disease status on demographic factors and b) regressing disease status on demographic factors and exposomic factors. Additionally, we ran sets of models where we constrained the space of exposomic factors to just MR-validated exposures in addition to separate sets of models considering a wider array of 196 exposures measured (albeit limited by number of complete cases), cumulatively leading to the specification of models A-I (Extended Data Table 5).

We estimated the variance (computed as Nagelkerke R^2^) explained by validated exposomic factors with demographic risk factors (Extended Data Table 6). Briefly, for each disease we run two major sets of logistic regression models: a) regressing disease status on demographic, genetic risk score (for CAD and T2D) and clinical (BMI for T2D) factors and b) regressing disease status on demographic factors and exposomic factors. Additionally, we ran sets of models where we constrained the space of exposomic factors to just MR-validated ones in addition to separate sets of models considering a wider array of 196 exposures measured (albeit limited by number of complete cases). Finally, we ran logistic regression models serially adding exposomic factors to the previous model in order of increasing FDR-corrected p-values until we ran the final model including the full wider array of exposomic factors (in addition to the baseline demographic and clinical risk factors). Additionally, pairwise correlations between categorical and numeric exposure variables were computed using the “polycor” R package.

## Code availability

We performed all analyses using R version 3.6.1. We made our code accessible at (https://github.com/stejat98/CardiometabolicExWASMR).

## Data availability

The data from the UK Biobank that support the findings of this study are available upon application (https://www.ukbiobank.ac.uk/register-apply/).

## Data Availability

All data produced are available in the Supplements. Individual-level participant are available by permission from the UK Biobank.

## Supplemental Table Legends

**Supplemental Table 1. Exposomic variables significant in ExWAS for incident CAD.** This table includes hazard ratios (HR), 95% confidence intervals (CI), FDR-corrected p-values and C-statistic from each cox proportional hazards model to systematically associate each exposure with time to event (CAD).

**Supplemental Table 2. Exposomic variables significant in ExWAS for incident T2D.** This table includes hazard ratios (HR), 95% confidence intervals (CI), FDR-corrected p-values and C-statistic from each cox proportional hazards model to systematically associate each exposure with time to event (T2D).

**Supplemental Table 3. Exposomic variables significant in ExWAS for prevalent CAD.** This table includes odds ratio (OR), 95% confidence intervals (CI), and FDR-corrected p-values from each logistic regression model to systematically associate each exposure with prevalent CAD.

**Supplemental Table 4. Exposomic variables significant in ExWAS for prevalent T2D.** This table includes odds ratio (OR), 95% confidence intervals (CI), and FDR-corrected p-values from each logistic regression model to systematically associate each exposure with prevalent T2D.

**Supplemental Table 5. Prevalent CAD versus incident CAD ExWAS significance classification table.** This table includes estimates of false positives, false negatives, true positives, true negatives when comparing prevalent CAD ExWAS (our “diagnostic test”) with incident CAD ExWAS findings. True Positives were ascertained as associations that had prevalent CAD and incident CAD effects concordant in direction in addition to FDR < 0.05 significance.

**Supplemental Table 6. Prevalent T2D versus incident T2D ExWAS significance classification table.** This table includes estimates of false positives, false negatives, true positives, true negatives when comparing prevalent T2D ExWAS (our “diagnostic test”) with incident T2D ExWAS findings. True Positives were ascertained as associations that had observational and MR beta estimates concordant in direction in addition to FDR < 0.05 significance.

**Supplemental Table 7. Exposomic variable associations for CAD from bi-directional two sample MR.** This table includes beta estimates and p-values for forward and reverse mendelian randomization (MR) analysis associating each exposure (using GWAS summary statistics derived from the UK Biobank) with CAD (using GWAS summary statistics derived from FinnGen).

**Supplemental Table 8. Exposomic variable associations for T2D from bi-directional two sample MR.** This table includes beta estimates and p-values for forward and reverse mendelian randomization (MR) analysis associating each exposure (using GWAS summary statistics derived from the UK Biobank) with T2D (using GWAS summary statistics derived from FinnGen).

**Supplemental Table 9. Exposomic variable associations for CAD across MR methods.** This table includes beta estimates and p-values for forward and reverse mendelian randomization (MR) analysis associating each exposure (using GWAS summary statistics derived from the UK Biobank) with CAD (using GWAS summary statistics derived from FinnGen) across the full suite of MR methods tested (MR Egger, weighted median, inverse variance weighted, simple mode, and weighted mode).

**Supplemental Table 10. Exposomic variable associations for T2D across MR methods.** This table includes beta estimates and p-values for forward and reverse mendelian randomization (MR) analysis associating each exposure (using GWAS summary statistics derived from the UK Biobank) with T2D (using GWAS summary statistics derived from FinnGen) across the full suite of MR methods tested (MR Egger, weighted median, inverse variance weighted, simple mode, and weighted mode).

**Supplemental Table 11. Observational versus MR significance classification table for lifestyle factors in association with CAD.** This table includes estimates of false positives, false negatives, true positives, true negatives when comparing ExWAS findings with respect to gold-standard causal estimates obtained from MR for CAD. True Positives were ascertained as associations that had observational and MR beta estimates concordant in direction in addition to the aforementioned observational and MR significance criteria.

**Supplemental Table 12. Observational versus MR significance classification table for lifestyle factors in association with T2D.** This table includes estimates of false positives, false negatives, true positives, true negatives when comparing ExWAS findings with respect to gold-standard causal estimates obtained from MR for T2D. True Positives were ascertained as associations that had observational and MR beta estimates concordant in direction in addition to the aforementioned observational and MR significance criteria.

**Supplemental Table 13. Observational versus MR significance classification table for social factors in association with CAD.** This table includes estimates of false positives, false negatives, true positives, true negatives when comparing ExWAS findings with respect to gold-standard causal estimates obtained from MR for CAD. True Positives were ascertained as associations that had observational and MR beta estimates concordant in direction in addition to the aforementioned observational and MR significance criteria.

**Supplemental Table 14. Observational versus MR significance classification table for social factors in association with T2D.** This table includes estimates of false positives, false negatives, true positives, true negatives when comparing ExWAS findings with respect to gold-standard causal estimates obtained from MR for T2D. True Positives were ascertained as associations that had observational and MR beta estimates concordant in direction in addition to the aforementioned observational and MR significance criteria.

**Supplemental Table 15. Effects of established clinical risk factors.** This table reports the odds ratios (OR) with 95% confidence intervals (CI) and p-values of established clinical risk factors from the literature including age, sex, family history, BMI, systolic blood pressure, diastolic blood pressure, HbA1c, LDL, HDL, triglycerides, total cholesterol, and smoking history.

**Supplemental Table 16. Ascertainment of Cardiometabolic Outcomes.** This table shows the ICD-10 and self-reported disease status codes used to identify coronary artery disease (CAD) and type 2 diabetes (T2D) disease status.

## Extended Data Table Legends

**Extended Data Table 1. Exposure variable breakdown.** This table shows the exposure categories as defined by Vermeulen et al.^3^ as well as the UKB, the number of variables within each category, the class of the variable category (i.e. whether the category contains biomarker (B), geographic (G) or questionnaire (Q) variables), and some example variables within each category.

**Extended Data Table 2. ExWAS-identified enrichment of exposomic categories (as defined by Vermeulen et al.**^3^**) for each disease.** This table shows the breakdown of the percentage of ExWAS-identified FDR < 0.05 associations by exposure categories as defined by Vermeulen et al for CAD and T2D.

**Extended Data Table 3. Observational versus MR significance classification table for CAD.** This table includes estimates of false positives, false negatives, true positives, true negatives when comparing ExWAS findings with respect to gold-standard causative estimates obtained from MR for CAD. True Positives were ascertained as associations that had observational and MR beta estimates concordant in direction in addition to the aforementioned observational and MR significance criteria.

**Extended Data Table 4. Observational versus MR significance classification table for T2D.** This table includes estimates of false positives, false negatives, true positives, true negatives when comparing ExWAS findings with respect to gold-standard causative estimates obtained from MR for T2D. True Positives were ascertained as associations that had observational and MR beta estimates concordant in direction in addition to the aforementioned observational and MR significance criteria.

**Extended Data Table 5. Breakdown of variance (R2) explained by exposomic and MR-validated exposomic factors with demographic risk factors.** This table contextualizes the variance (computed as R^2^) explained by exposomic, MR-validated exposomic factors with demographic risk factors. Delta R^2^ is computed as the difference between R^2^ computed from full model and R^2^ computed model only including demographic and clinical risk factor covariates.

**Extended Data Table 6. Baseline demographic breakdown of samples used for models A-I.** This table shows the breakdown of the mean and shows the mean or percentage of key demographic variables for the samples of White UKB participants that were primarily used for our models A-I including age, sex, average household income, and assessment center.

